# Assessing the variations in breast/ovarian cancer risk for Chinese BRCA1/2 carriers

**DOI:** 10.1101/2020.07.20.20135202

**Authors:** Ang Li, Yi Zi, Jiaqi Luo, Xiaobin You, Zhaoji Lan, Tianliangwen Zhou, Yangming Wu, Qihuan Zhi, Huijun Su, Mei Zhu, Siwen Xu, Yun Gao, Zaixuan Zhong, Ling Xie, Yuanqin Wang, Qiuping Lin, Xiaoting Li, Jiamin Zhan, Hui Weng, Dan Li, Shulan Xu, Gang Sun, Yujian Shi

## Abstract

**Background:** Cancer risks vary in different BRCA1/2 mutations. Previous studies based on Caucasian population have identified regions associated with elevated/reduced risks of breast/ovarian cancers. Since ethnic differences are known to affect BRCA1/2 mutation spectra, we are interested in defining Chinese-specific ovarian/breast cancer cluster regions (OCCR/BCCR) and comparing with previously reported Caucasian-based cluster regions. We also aim to characterize the distribution and estimate the cancer risks of different Chinese recurrent mutations.

**Methods:** 7,919 (3,641 unselected cancer-free women + 4,278 female cancer patients) individuals were included in the study. Germline BRCA1/2 status were detected with amplicon-based next-generation sequencing. BRCA1/2 carriers were defined as bearing likely pathogenic or pathogenic mutations. We calculated odds ratio (OR) of breast cancer and OR of ovarian cancer, and their ratio of the two ORs (ROR) for each region. ROR > 1 indicated elevated odds of breast cancer and/or decreasing odds of ovarian cancer; ROR < 1 indicated increasing odds of ovarian cancer and/or decreasing breast cancer odds. The frequency, distribution and penetrance of six known Chinese founder mutations were characterize respectively. Haplotype analysis and age estimation were performed on the most prevalent and widely-spread founder mutation BRCA1:c.5470_5477del.

**Results:** A total of 729 subjects were detected with germline BRCA1/2 deleterious mutations, including 236 BRCA1 and 122 BRCA2 mutations. The putative Chinese OCCR/BCCR are partially overlapped with Caucasian-based OCCR/BCCR and shared structural-functional characteristics. The six known Chinese founder mutations vary greatly in both distribution and penetrance. The two most prevalent and widely-spread mutations are estimated to convey low penetrance, while the area-restricted founder mutations seemed to confer higher or nearly complete penetrance. The most prevalent founder mutation BRCA1:c.5470_5477del accounting for 9.5% - 18% of BRCA1 carriers is estimated to have emerged ∼2,090 years ago (70 B.C.) during the Han Dynasty, about 290 years (∼14.5 generations) prior to the Three Kingdoms Period when a major population migration occurred.

**Conclusion:** BRCA1/2 carriers with different genotypes have significantly different cancer risks. Hence ideally risk assessment should be mutation-specific, rather than concerning a single figure. The probably most ancient Chinese founder mutation may have originated more than 2,000 years ago.

## Background

Since the establishment of predisposing effects of BRCA1 (MIM:113705) and BRCA2 (MIM: 114480) to breast/ovarian cancers, we have accumulated understandings of the biological roles of BRCA1/2 and their prevalence of mutations in different ethnic populations through a lot of functional experiments and sample-based studies. It is now generally believed that deleterious mutations in BRCA1/2 or other related genes result in production of malformed proteins that are unable to function during the error-free DNA repair process mediated by homologous recombination (HR) upon DNA double-strand break, which will lead to genomic instabilities and eventually the development of malignancies. It is also known that race/ethnic differences are presented in mutation spectrum, prevalence of mutations and in recurrently mutated positions which are likely to reflect the so called “founder effects”. For example, Ashkenazi Jewish generally have higher risk of being BRCA1/2 carriers because of the highly prevalent founder mutations BRCA 185delAG, BRCA1 5382insC and BRCA2 6174delT [1]. Several Chinese founder mutations have also been reported previously [2–6]. Moreover, it is recognized that great differences in cancer risks even present within BRCA1/2 mutation carriers, depending on the location and type of mutation they bear. According to previous observations, mutations within certain regions (defined as ovarian cancer cluster regions, OCCRs) are associated with higher ovarian cancer and/or lower breast cancer risks than other regions, and certain regions (defined as breast cancer cluster regions, BCCRs) with higher breast cancer and/or lower ovarian cancer risks. The biological impact caused by the mutation is the determining factor of cancer risk, which apparently affects the position of the OCCRs and BCCRs; the race/ethnicity of the studied population is another variable as it affects the mutation spectrum (e.g. the position of mutation hotspots). Several studies have calculated putative OCCR/BCCRs of BRCA1/2 using samples mostly of Caucasian origin, and the estimated regions are considerably overlapping. Interestingly, for both BRCA1 and BRCA2, the estimated OCCRs seemed to locate in the centre of the CDS and significantly overlap with the largest exon (EX10 of BRCA1 and EX11 of BRCA2), while the BCCRs seemed to occupy the 5’ and the 3’ ends of the CDS. As we expect race/ethnic difference to cause some degree of variability in OCCR/BCCRs when studying different populations, and since Asian populations only represented 1% of the total samples in previous studies, we are interested in defining OCCR/BCCRs in Chinese BRCA1/2 carriers and finding out to what extent the position of OCCR/BCCRs could be varied. Moreover, it has been widely acknowledged that the identification and screening of founder mutations are highly cost-effective measures for cancer risk management [7]. Hence we also aim to characterize the distribution of Chinese recurrent mutations (or founder mutations) and estimate the cancer risks they confer, in order to provide a reference to the precision management of genetic risks for Chinese BRCA1/2 carriers.

## Methods

### Samples

A total of 3,641 unselected cancer-free women and 4,278 female cancer patients (breast cancer, ovarian cancer, colon cancers and pancreatic adenocarcinoma) in the nationwide of China volunteered to enroll in this study. Clinical characteristics of samples in relation to BRCA1/2 are presented in Table 1. Among the 3,641 cancer-free individuals, 2,615 were negative of family history and further selected as healthy control for penetrance estimation and haplotype analysis. All the subjects involved had given their written informed consent in accordance with the Chinese ethical standards and the 2008 Helsinki declaration.

**Table 1.** Clinical characteristics and germline BRCA1/2 status of the samples in this study.

| Cancer site | Number, n=7919(%) | Median age | P-value | BRCA carriers, n=752(%) | Non-carriers, n=7190(%) | P-value |
| --- | --- | --- | --- | --- | --- | --- |
| Breast | 2400(30.31%) | 45.00(38.00-52.00) | <0.0001 | 285(11.88%) | 2115(88.13%) | 0.041 |
| Ovary | 1679(21.20%) | 52.00(46.00-59.00) |  | 332(19.32%) | 1365(80.68%) |  |
| Breast and ovary | 32(0.40%) | 55.00(48.25-60.75) |  | 16(48.48%) | 16(51.52%) |  |
| Breast/ovary and | 33(0.42%) | 53.50(45.00-61.00) |  | 1(3.03%) | 32(96.97%) |  |
| Other sites | 116(1.46%) | 61.00(49.25-70.75) |  | 5(4.31%) | 111(95.69%) |  |
| Cancer-free | 3641(45.98%) | 36.00(30.00-43.00) |  | 90(2.47%) | 3551(97.53%) |  |

### BRCA1/2 genetic testing and mutation classification

Blood samples were collected from all enrolled individuals and subjected to NGS-based BRCA1/2 whole-exon sequencing (all coding regions and exon-intron boundaries ± 20bp). The analysis pipeline and mutation classification followed protocols as described in our previous research [8]. The human genome hg19/GRC37 was used as reference; the NCBI reference sequences NM_007294.3 and NM_000059.3 were used for annotations of BRCA1 and BRCA2 variants respectively. Nomenclature of mutations followed the latest version of Human Genome Variation Society Sequence Variant Nomenclature (HGVS, http://varnomen.hgvs.org/) and Mutalyzer Name Checker (http://mutalyzer.nl). Deleterious mutations in this study include likely pathogenic and pathogenic mutations. Further validations of all deleterious mutations and variants of uncertain significance were performed by Sanger sequencing. Visualization of the variants were presented with Circos plots [9].

### Mutation grouping and statistical analysis

To estimate OCCR/BCCR, segments of regions containing all deleterious mutations (regardless of mutation type or function) need to be created for statistical calculations. We divided the coding sequence (CDS) of BRCA1 and BRCA2 into bins by base pair location so that each bin contains roughly equal number of carriers (Table 2). Large rearrangements were excluded to avoid spanning multiple bins.

**Table 2.**
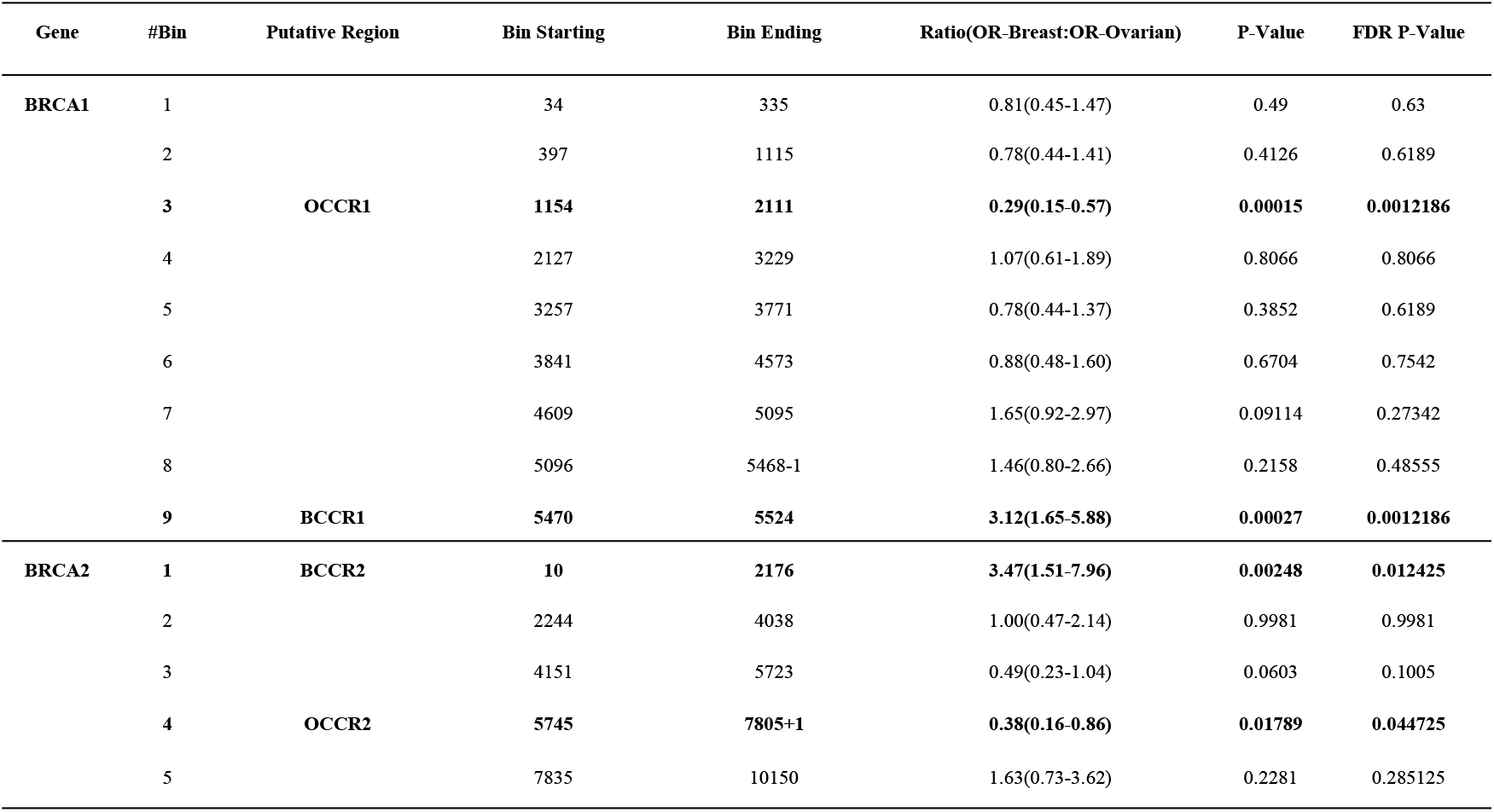
Ovarian cancer cluster regions (OCCR) and breast cancer cluster regions (BCCR) in BRCA1 and BRCA2.

We then calculate the odds ratio (OR) of breast cancer and the OR of ovarian cancer respectively for each bin. We computed a statistical measure ROR, defined as the ratio of breast versus ovarian cancer OR. The value of ROR is associated with elevated/reduced breast or ovarian cancer odds (ROR > 1, increasing odds of breast cancer and/or decreasing odds of ovarian cancer; ROR < 1, increasing odds of ovarian cancer and/or decreasing breast cancer odds). ROR of all bins were compared to identify significant outliers as putative OCCR/BCCRs. P-values were adjusted with Benjamini-Hochberg procedure to reduce the possibility of false positive.

### Characterization of BRCA1:c.5470_5477delATTGGGCA

Among the recurrent mutations identified in this study, BRCA1:c.5470_5477delATTGGGCA occurred in 9.5% (50/527) of BRCA1 carriers and had enough materials for further experimental validation. We then performed haplotype analysis on this variant. A total of 81 subjects (31 patients with the same mutations from independent families, 50 unrelated controls without the mutation were included in the haplotype analysis. According to marker selection methods described previously [4,10,11], we selected nine polymorphic markers flanking the BRCA1 gene - D17S800, D17S1320, D17S1321, D17S855, D17S1323, D17S1327, D17S1326, D17S1325, D17S791 (short tandem repeats, STR, spanning approximately 5.8Mbp, supplementary Figure S1). We detected STR lengths using fluorescently end-labeled PCR primers and ABI 3730xl Genetic Analyzer (Applied Biosystems, Foster City, CA, USA). We reconstructed all possible haplotypes by using PHASE v.2.1.1 [12] (supplementary Table S1). Chi-square test was used to evaluate the differences in allele frequencies between patients and healthy subjects for each STR involved.

We also performed age estimation of BRCA1:c.5470_5477delATTGGGCA. We used DMLE+ v2.3 to estimate the original time when the mutation emerged in the BRCA1 gene [13]. Based on a Markov chain Monte Carlo algorithm, this program enabled Bayesian inference to estimate the age of the specific mutation with the knowledge of the patients’ and controls’ haplotypes or genotypes observed, physical distances between markers (Mb) and the estimated population growth rate. The population statistics of ancient time points were taken from historical data [14]; population figure of year 1949 was collected from the National Bureau of Statistics of China (http://data.stats.gov.cn/index.htm).

### Penetrance estimations on recurrent mutations

We estimated the penetrance of the recurrent mutations using the allelic model by Bayes’ theorem [15]. This formula required the knowledge of the lifetime risk of breast/ovarian cancer of Chinese population. Restricted by the access to the official data, we used the estimated mean value of lifetime risk (0.053) for breast cancer derived from the Gail Model among Chinese women [16] as an approximate replacement.

## Results

Out of the total 7,919 samples containing 4,278 cancer patients and 3,641 cancer-free individuals, 729 were detected with germline BRCA1/2 deleterious mutations. Median age of breast cancer diagnosis was 43, 44 and 46 in BRCA1 carriers, BRCA2 carriers and non-carriers; median age of ovarian cancer diagnosis was 51, 54 and 52 in BRCA1, BRCA2 carriers and non-carriers, respectively.

A total of 236 deleterious germline mutations in BRCA1 (Figure 1) and 122 in BRCA2 (Figure 2) were detected and verified in this study. Several recurrent mutations are identified (Figure 1&2, highlighted with red, bigger font). BRCA1:c.5470_5477delATTGGGCA (Ile1824AspfsTer3) was the top hit, accounting for 9.5% (50/527) BRCA1 carriers; the second hit for BRCA1 was c.981_982delAT, with 4.0% (21/527) prevalence in BRCA1 carriers; next were c.3770_3771delAG (12/527, 2.3% of BRCA1 carriers), c.5521delA (11/527, 2.1%) and c.4801A>T (8/527, 1.5%). The BRCA2:c.5722_5723delCT was the top hit for BRCA2 (10/202, 5.0%), followed by BRCA2:c.3109C>T (9/202, 4.5%).

**Figure 1.**
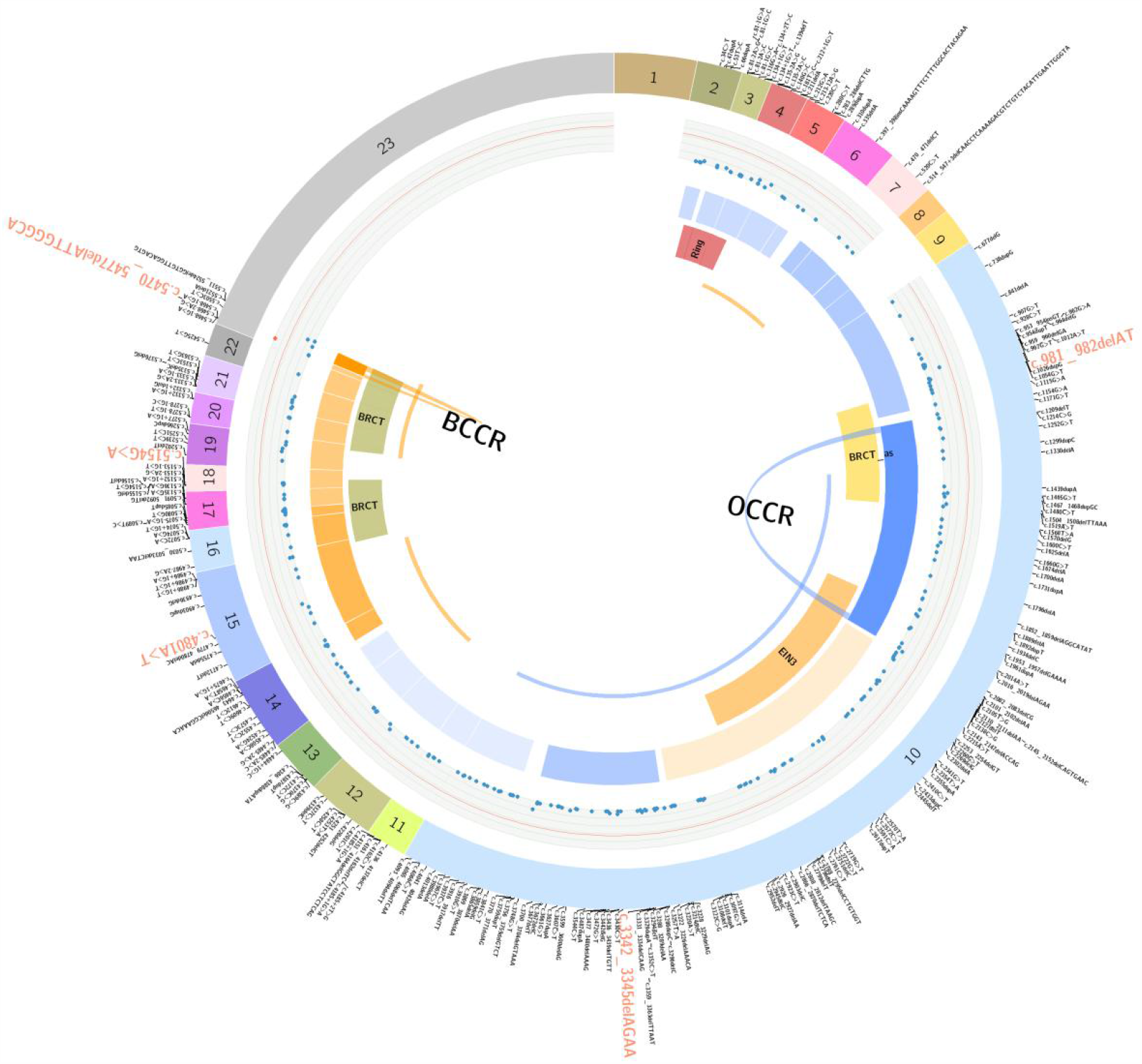
Circos plot representation of all BRCA1 deleterious mutations identified in this study. The outmost ring displays the 23 exons of BRCA1 and each variant at corresponding positions. Recurrent variants are highlighted in red and enlarged font. The second circle is the mutation density graph, each dot correspond to a variant, and its distance to the outmost ring represents the frequency (the closer the higher; the most prevalent mutation is colored in red). The ROR is represented by heatmap in the intermediate ring (ROR>1, in orange; 0<ROR<1, in blue). Next circle displays the BRCA1 functional domains. The innermost arcs represent previously reported Caucasian-based OCCR/BCCRs. The areas enclosed by the arches indicate the estimated Chinese OCCR/BCCR, which are statistically significant (FDR p-value<0.05).

**Figure 2.**
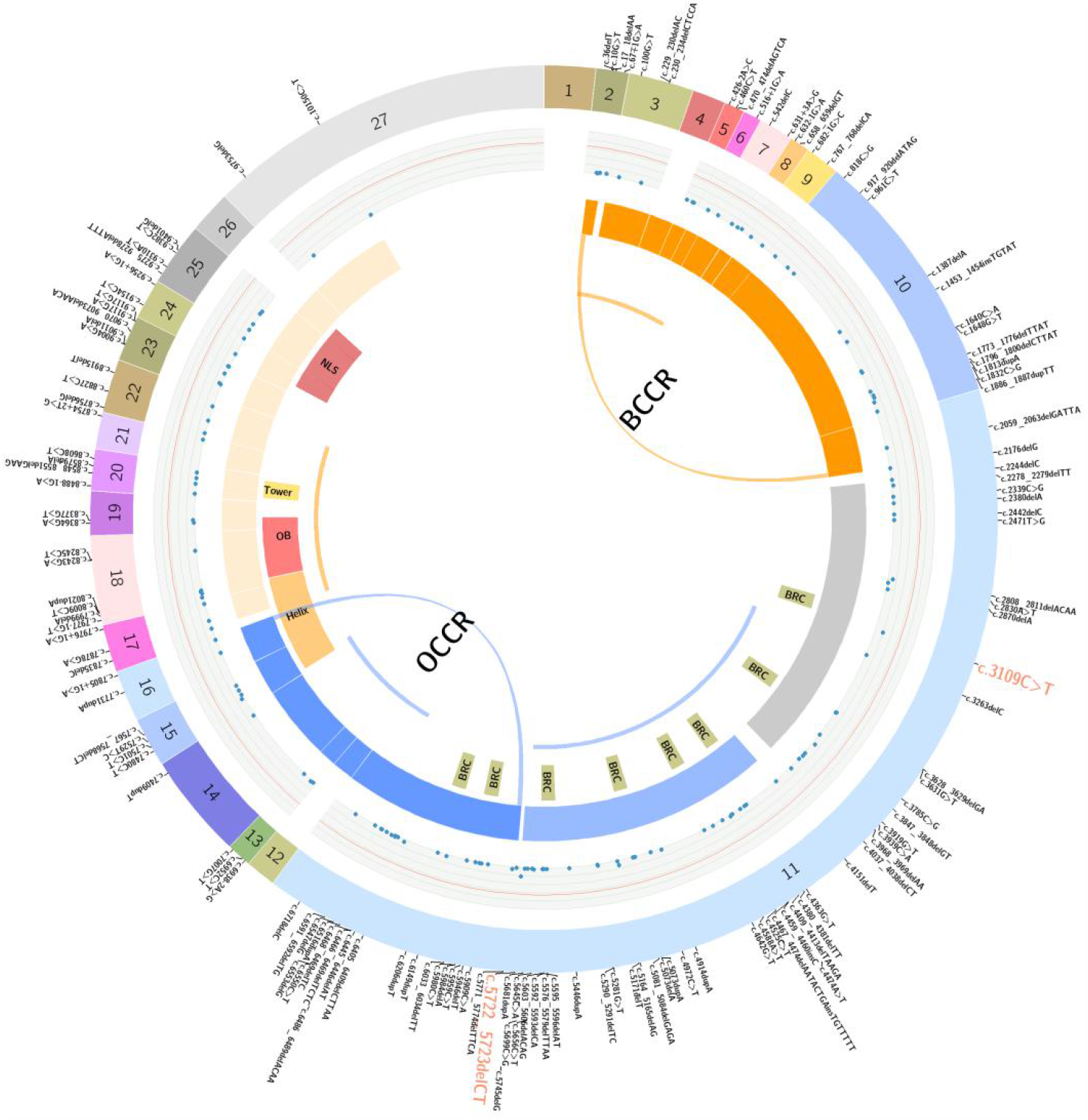
Circos plot representation of all BRCA2 deleterious mutations identified in this study. The outmost ring displays the 27 exons of BRCA2 and each variant at corresponding positions. Recurrent variants are highlighted in red and enlarged font. The second circle is the mutation density graph, each dot correspond to a variant, and its distance to the outmost ring represents the frequency (the closer the higher; the most prevalent mutation is colored in red). The ROR is represented by heatmap in the intermediate ring (ROR>1, in orange; 0<ROR<1, in blue). Next circle displays the BRCA2 functional domains. The innermost arcs represent previously reported Caucasian-based OCCR/BCCRs. The areas enclosed by the arches indicate the estimated Chinese OCCR/BCCR, which are statistically significant (FDR p-value<0.05).

### Estimated Chinese OCCR/BCCR in BRCA1

We predicted an OCCR at c.1154 - c.2111 (Table 2; Figure 1, blue arch) with a relative decrease in breast cancer risk and a relative increase in ovarian cancer risk (ROR = 0.29; 95% CI = 0.15 - 0.57; FDR-corrected p-value = 1.2 × 10^−3^). The putative OCCR lies within the largest exon (Exon 10) of BRCA1 and is partially overlapped with previously reported OCCR (c.1380 - c.4062)[17] (Figure 1, blue arc). The putative OCCR entirely or partially spans several functional domains, including the binding sites for Rb, Rad50, c-Myc and the nuclear localization sequence (NLS) [18]. A putative BCCR was found at c.5470 - c.5524 near the 3’ end of the CDS (Table 2; Figure 1, orange arch), explained by a relative decrease in ovarian cancer risk (ROR = 3.12, 95% CI = 1.56 - 5.88, FDR-corrected p = 1.2 × 10^−3^). The BCCR lies within the second BRCT domain (c.5268 - c.5526) and is also partially overlapped with previously reported BCCR (c.5261 - c.5563)[17] (Figure 1, orange arc).

### Estimated Chinese OCCR/BCCR in BRCA2

We predicted an OCCR at c.5745 - c.7805+1 (Table 2; Figure 2, blue arch) with a relative decrease in breast cancer risk and a relative increase in ovarian cancer risk (ROR = 0.38; 95% CI = 0.16 - 0.86; FDR-corrected p-value = 0.044). The putative OCCR is partially overlapped with previously reported OCCR (c.6645 - c.7471)[17] (Figure 2, blue arc), spanning from the last two BRC repeats within Exon 11 (the largest exon of BRCA2) to approximately the boundary of Exon 17 and 18, which contains the most part of the helical DNA binding domain (c.7437 - c.8001). A putative BCCR was found at c.10 - c.2176 (Table 2; Figure 2, orange arch), explained by a relative increase in breast cancer risk and a relative decrease in ovarian cancer risk (ROR = 3.47, 95% CI = 1.51 - 7.96, FDR-corrected p = 0.012); it is also partially overlapped with previously reported BCCR (c.1 - c.596, c.772 - c.1806)[17] (Figure 2, orange arcs). The putative BCCR spans from the 5’ end of the CDS to approximately the boundary of Exon 10 and 11.

### Penetrance vary among different Chinese founder mutations

We estimated the breast/ovarian cancer penetrance of six previously reported founder mutations based on the clinical data of our samples (see Materials and Methods for details). Here we used the estimated population lifetime risk of breast cancer for Chinese women derived from the Gail Model [16] (about 5.3%; Figure 3, gray vertical line) as the population baseline risk, and the approximate lower bound (0.35; Figure 3, black dashed line) of breast/ovarian cancer risk in BRCA1/2 carriers (according to previous studies [19,20]) as an minimum expected penetrance of all deleterious mutations throughout the whole gene. Of the six founder mutations examined, the two most prevalent mutations (BRCA1:c.5470_5477del and BRCA1:c.981_982del) were estimated to have penetrance lower than 0.35; one (BRCA2:c.3109C>T) with either above or lower than 0.35 penetrance subjected to family history (high penetrance only occurs in carriers with positive family history); and three with small sample sizes (BRCA1:c.3342_3345del, BRCA1:c.5154G>A, BRCA1:c.4801A>T) showed 100% complete penetrance due to no carriers found in control. Our results demonstrated that the penetrance of different BRCA1/2 deleterious mutations vary greatly and show large deviation from the expected value.

**Figure 3.**
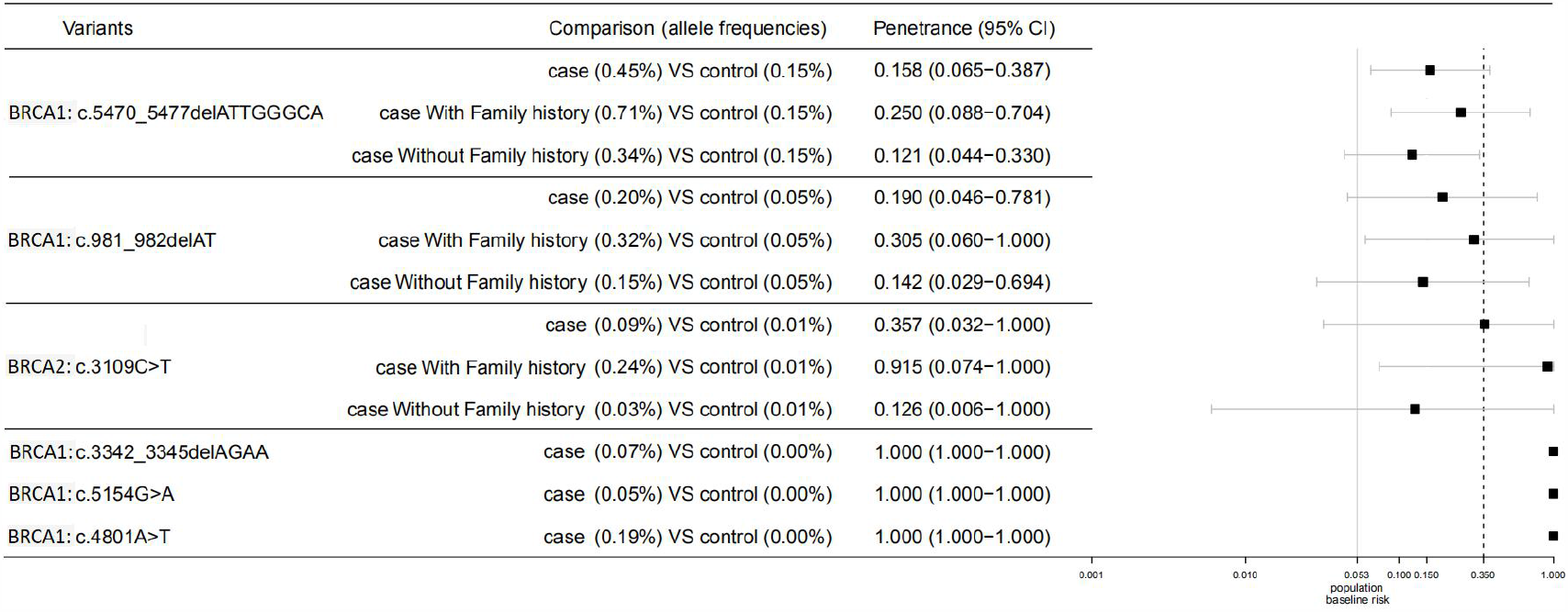
Penetrance estimations of six known Chinese founder mutations. The estimated population lifetime risk of breast cancer for Chinese women (derived from the Gail Model) is 0.053; the estimated breast/ovarian cancer risk in BRCA1/2 carriers is 35-50% (lower bound: 0.35). Great variations in penetrance are observed among the six founder mutations.

### The founder BRCA1:c.5470_5477del is estimated to have emerged more than 2000 years ago

As mentioned above, the BRCA1:c.5470_5477del was the most recurrent deleterious mutation, accounting for 9.5% of BRCA1 mutation carriers. Haplotype analysis was carried out on 31 unrelated patients and 50 unrelated controls without the mutation. The haplotype analysis was performed independent of the work of Meng et al. [6]; similar to their findings, our haplotype analysis suggested strong founder effect (supplementaryTable S1) of the mutation. Moreover, carriers of this variant are distributed throughout the country (Figure 4), except provinces with sampling size < 100 (regions colored in gray; total sample size: 7919). Compared with other known Chinese founder mutations, BRCA1:c.5470_5477del (orange dots) showed the highest allele frequency and the most thorough spread in terms of geographical location, indicating a relatively early emergence of the mutation. To further estimate its possible time of emergence, we used the formula shown below to calculate a list of average growth rates (‰ per year) from official population records [14] of different historical time points ranging from 684 B.C. to 1949 A.D. (Table 3). The calculated growth rates differ slightly to each other, with a maximum of 7.81‰(1741 A.D. - present) and a minimum of 1.54‰(2 A.D. - present). For reference, the estimated growth rate of year 2019 is 3.34‰ (http://data.stats.gov.cn/index.htm).

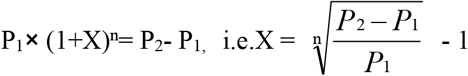

X = average growth rate, n = time till now (years),

P_1_ = population as recorded n years ago, P_2_ = Chinese population in 2020 = 1.4 billion

**Table 3.** Age estimation analysis using population data from different historical time points.

| Time point | Population (thousand) | Average growth rate | Time till now (yrs) | Estimated mutation age (yrs) |
| --- | --- | --- | --- | --- |
| 684 B.C. | 11,840 | 1.76‰ | 2,704 | 2,145 |
| 2 A.D. | 59,590 | 1.54‰ | 2,018 | 2,090 |
| 609 A.D. | 46,020 | 2.40‰ | 1,411 | 1,534 |
| 1110 A.D. | 46,730 | 3.71‰ | 910 | 1,585 |
| 1403 A.D. | 66,600 | 4.87‰ | 617 | 992 |
| 1741 A.D. | 143,410 | 7.81‰ | 279 | 602 |
| 1949 A.D. | 541,670 | 6.50‰ | 71 | 1,006 |

**Figure 4.**
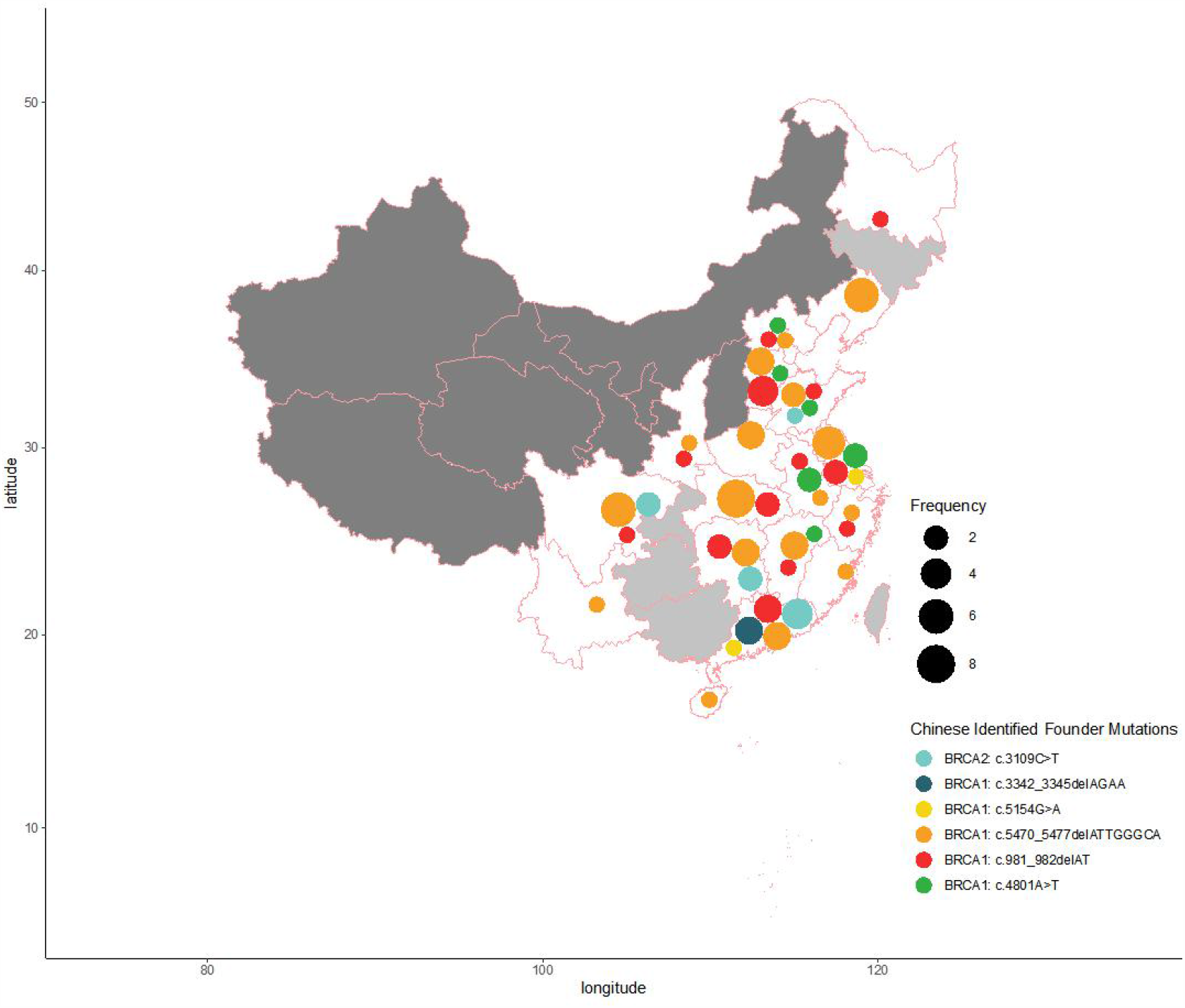
Geographical distribution of the six known Chinese founder mutations. Areas with sample size < 100 are colored in light gray; areas with sample size < 50 are colored in dark gray. Total sample size is 7,919. Spot size indicates frequency of mutation, spot color indicates different founder mutations.

We used DMLE+2.3 to estimate the distribution of possible mutation age (years) under each growth rate (supplementary Figure S2), then we compared the estimated mutation age (peak) and the actual time from which the growth rate is drawn. Among the seven growth rates used, the estimated mutation age (n) and the actual time (n) showed the best correlation with 1.54‰(2 A.D. - present), for which n=2090, n=2018. The estimated time (∼2,090 years ago, i.e. ∼70 B.C.) of emergence of the mutation lies within the period of Han Dynasty (206 B.C. - 220 A.D.), currently known as the second imperial dynasty of China.

## Discussion

We have presented a large-scale BRCA1/2 screening in both cancer patients and cancer-free individuals throughout China. We preliminarily defined the Chinese OCCR/BCCRs in BRCA1/2 and compared with the previously reported OCCR/BCCRs based on mainly Caucasian population. We estimated penetrance for each of the six known Chinese founder mutations and demonstrated great variations between them. Unlike previous studies that based on patients and/or their family members, this study included a large number of samples from the normal population, allowing us to assess the risk of a mutation carrier without family history, which can significantly differ from the risk of those with family history. We also performed haplotype analysis on the most recurrent founder mutation BRCA1:c.5470_5477del and estimated its time of emergence to be ∼2,090 years ago within the Han Dynasty.

Our OCCR/BCCRs partially overlap with previously reported OCCR/BCCRs drawn from white/Jewish population [17]. We observed both in the Chinese and the white/Jewish studies that OCCRs position at the middle of the gene where the largest exon with the highest mutation rate is located, and with binding sites for key proteins involved in DNA repair processes such as RAD51 and PALB2 [18,21]. Whereas BCCRs tend to position at the ends (5’ and/or 3’) of the gene where smaller exons with secondary peaks of mutation rate are located, and include transcription activation domain (TAD) and DNA-binding domain (DBD). OCCRs tend to be longer than BCCRs. The location of OCCR/BCCRs is thought to be determined mainly by two factors: the biological impact caused by mutations within a certain region, and the frequency of these mutations detected within the studied population. While the former is likely not to be affected by ethnicity and may explain the concordant parts of the OCCR/BCCRs, the latter is known to be considerably affected by ethnicity and may explain the parts with discordance (38% of Chinese BRCA1/2 variants have not been reported in other populations [22]). Indeed, the Caucasian OCCR/BCCRs and Chinese OCCR/BCCRs show concordance with hotspot exons (i.e. frequently mutated exons) drawn from BIC and dbBRCA-Chinese databases [22], respectively. The difference in mutation frequency of each exon between the two databases may explain some of the difference shown between our Chinese OCCR/BCCRs and the previously reported Caucasian-based OCCR/BCCRs. For example, the reported mutation rate of BRCA1 Exon5 in BIC is 3-fold of that in dbBRCA-Chinese database, which may explain the presence of a BCCR in Caucasian data and its absence in our Chinese data (note that due to historical reasons, the first exon was named Exon2 so that Exon11 is the largest exon, as seen in many literature and databases; and that Exon2,5,11 are equal to Exon1,4,10 in our study). However, the exact mechanism of how the biological impact is affected by the position of the mutation is unclear, since the vast majority of BRCA2 mutations are truncating and able to trigger nonsense-mediated mRNA decay (NMD) [23]; truncated BRCA2 proteins are cytoplasmic [24] and unable to enter the nucleus due to lack of an intact nuclear localization signal. One possible explanation might be that NMD is not removing all truncating-transcripts, which translate into truncated proteins and might compete with intact proteins for binding partners at the cytoplasm, or even become able to enter the nucleus through the carriage by a binding partner.

With the awareness of the great heterogeneity in cancer risks for mutations in different regions as evidenced by the OCCR/BCCRs, we focused on estimating independent penetrance for each recurrent mutation instead of a single penetrance for the whole gene. The two most prevalent mutations which have been shown to spread through the country are estimated to have a relatively low penetrance, while those less prevalent ones residing in a local scope tend to have higher or complete penetrance. Our results demonstrated that “one risk does not fit all”, as suggested previously by De Bock et al. [25]. Not every BRCA1/2 carrier has the same risk of developing cancers; some may never develop cancer throughout their lives. It is therefore important to consider separately when assessing cancer risks for BRCA1/2 carriers with different genotypes. While a regular ultrasound check for carriers with low risk genotype would be enough, a more preventive strategy such as risk-reducing resection should be considered for carriers with high-risk genotype [26]. Furthermore, assisted reproductive technology [27] should be considered for carriers who are in childbearing age and with high risk genotype to prevent the passage of the high-risk allele. For those putative high-risk mutations that seemed to be limited within a relatively small local area, it is necessary to carry out concentrated screening in order to further verify the penetrance, and if the high penetrance is confirmed, to identify more carriers of the mutation and take early preventative measures.

We estimated the time of emergence of currently the most recurrent Chinese founder mutation (also the most widely spread and probably the oldest), BRCA1:c.5470_5477del. Our estimation indicated that the emergence of this mutation may have happened ∼2,090 years ago (∼70 B.C.) during the Han Dynasty, or more specifically, during the Western Han (Xi Han, 206 B.C. - 8 A.D.). The thorough spread of the mutant allele throughout the country is most likely to be explained by multiple large-scale population migration events caused by frequent wars. There have been three major waves of population migration in Chinese history: the first started from the Three Kingdoms Period, i.e. the end of the Han Dynasty (220 A.D.), until the end of the Southern and Northern Dynasties (also known as the start of the Sui Dynasty, 589 A.D.); the second occurred during the An-shi Rebellion in Tang Dynasty (755 A.D.); the third happened during the Jingkang Incident (1127 A.D.) which has led to the end of the Northern Song Dynasty. The three major migrations all happened after the estimated time of emergence of BRCA1:c.5470_5477del. There are 290 years (∼14.5 generation) between the estimated emergence (70 B.C.) and the first wave of migration (220 A.D.), which would be enough for the initial accumulation of mutant alleles so that the founder mutation can survive wars and natural disasters, and inherit for more than two thousand years.

There are several limitations of this study. It is retrospective and the number of subjects may be insufficient for a comprehensive estimation of the 1.4 billion Chinese population; follow-up time for BRCA1/2 carriers are short, and most of the cancer-free carriers are under 70 years old, which can cause underestimation of the cancer risk; the portion of BRCA1/2 carriers in cancer-free individuals is higher than expected, which is likely due to the fact that those with family histories are more willing to participate in testing; we did not include large rearrangement events of BRCA1/2 in our research, which may account for more than 5% of all BRCA1/2 mutations [28]; due to the lack of genome-wide single-nucleotide polymorphism of all subjects included, we have no access to the population stratification considering the geographical difference and 56 ethnic-origins of Chinese population.

In conclusion, we preliminarily defined the OCCR/BCCRs based on a large number of Chinese samples. The Chinese OCCR/BCCRs partially overlap with the previously defined OCCR/BCCRs based on Caucasian samples. We estimated the penetrance of the six major Chinese founder mutations respectively and demonstrated great variations between them, which strongly suggests cancer risks should be calculated and considered separately depending on the genotype rather than looking at a fixed risk figure. Finally, we investigated the most prevalent and nationally-spread Chinese founder mutation BRCA1:c.5470_5477del, and estimated that it has more than two thousand years of history.

## Data Availability

Supplementary figures and tables are provided in supplementary data. Mutational data can be made available upon reasonable request.

## Declarations

### Ethics Approval and Consent for Participation and Publication

All procedures performed within this study were done in accordance with the Chinese ethical standards and with the 2008 Helsinki declaration.Informed written consent for participation and publication was obtained from each participant.

## Acknowledgements

The authors would like to thank all participants of this study for their contributions to scientific research.

## Conflict of Interest

Ang Li, Yi Zi, Yangming Wu, Xiaoting Li, Jiamin Zhan were employed by Top Gene Tech (Guangzhou) Co., Ltd. when the work was conducted, but no longer by the time of manuscript submission. Other authors declare no competing interests.

## Notes

### Funding Statement

None to be declared.

### Author Declarations

This study was approved by the Ethics Committee of Daping Hospital, Army Medical University. All procedures performed within this study were done in accordance with the Chinese ethical standards and with the 2008 Helsinki declaration.Informed written consent for participation and publication was obtained from each participant.

